# Factors associated with ocular injury among patients admitted with head injury at Mulago National Referral Hospital a tertiary hospital in Uganda

**DOI:** 10.64898/2026.08.18.26360753

**Authors:** Alfred Olupot, Faith Oguttu, Joshua Shiuma, Oscar Jude Lyazzi, Bernard Odong, Mlaluko Rajabu Jumanne, Kamya Frank, Jacob Ntende, David Mukunya, Juliet Otiti-Sengeri, Anne Ampaire Musika, Rebecca Claire Lusobya, Ssali Grace Nsibirwa, Immaculate Atukunda

## Abstract

**Background:** Given the close anatomical proximity of the eye and its adnexae to the cranium, ocular injury frequently coexists with head trauma. However, specific factors predisposing patients with head injury to ocular involvement in Uganda remain poorly defined.

**Objective:** This study aimed to determine the factors associated with ocular injury among patients with head injury at Mulago National Referral Hospital (MNRH), a tertiary referral facility in Kampala, Uganda.

**Methods:** We conducted a cross-sectional study among 383 adult patients admitted with head injury to the Accidents and Emergency Department of MNRH from 15/05/2025 to 30/07/2025. All participants underwent a standardized ophthalmic evaluation. Bivariable and multivariable Poisson regression analyses were used to identify factors associated with ocular injury, reported as adjusted prevalence ratios (APR).

**Results:** Ocular injury was identified in 268 of 383 patients (70.0%; 95% CI: 65.1-74.5). On multivariable analysis, age 26-44 years (APR 1.18; 95% CI: 1.11-1.43; p=0.013), commercial motorcyclist (boda-boda rider) occupation (APR 1.25; 95% CI: 1.06-1.47; p=0.007), road traffic accident mechanism of injury (APR 1.19; 95% CI: 1.02-1.38; p=0.03), severe head injury (APR 1.70; 95% CI: 1.44-2.01; p<0.0001), and the presence of facial fractures (APR 1.61; 95% CI : 1.44-1.81; p<0.0001) significantly increased the likelihood of ocular injury. Conversely, intracranial hemorrhagic lesions were inversely associated with ocular injury (APR 0.80; 95% CI: 0.68-0.93; p=0.004).

**Conclusion:** Ocular injuries are common among patients with head injury. Patients aged 26 to 44 years, commercial motorcyclist occupation, road traffic injury, severe head injury, and facial fractures have a higher risk of ocular injury. We recommend prioritizing ophthalmic evaluation of for patients admitted with head injury to minimize preventable visual impairment.

## Introduction

In Uganda and across Sub-Saharan Africa, head injury ranks among the leading causes of trauma-related morbidity and mortality [1, 2]. At Mulago National Referral Hospital (MNRH), approximately 95 head injury patients are admitted each month, most following road traffic collisions [3, 4]. Due to the structural continuity between the orbit, cranium, and mid-facial skeleton, mechanical forces applied to the cranium readily extend into the visual system, resulting in a high incidence of concurrent ocular trauma [5]. Globally, an estimated 60 million ocular injuries occur each year, and low- and middle-income countries (LMICs) account for roughly 60% of this burden [6, 7]. Compared with patients admitted for other categories of concomitant trauma, those with head injury carry a threefold higher likelihood of a concurrent ocular injury [8, 9]. Nevertheless, concurrent ocular trauma is easy to miss in the acute setting because an obtunded patient cannot fully cooperate with proper eye examination, swollen eye lids obscure the globe, and clinical attention is understandably directed toward stabilizing the patient’s airway, breathing, circulation and neurological status first [10]. Since delayed recognition of some of these ocular injuries can translate into avoidable visual impairment or blindness, early identification matters [10]. However, what predisposes head injury patients to concurrent ocular trauma in this setting remains unclear. Earlier studies at MNRH focused exclusively on road crash casualties [11] or lacked a comprehensive ophthalmic examination protocol [12], leaving the combined influence of socio-demographic and clinical factors not fully examined. Establishing these risk factors helps clinicians to prioritize high-risk patients for ophthalmic review and reduces preventable visual loss. Consequently, this study aimed to determine the factors associated with ocular injury among head injury patients admitted to MNRH in Kampala, Uganda.

## Materials and methods

### Study design and setting

This was a hospital-based, analytical cross-sectional study conducted to identify the socio-demographic and injury-related correlates of ocular injury among adult head trauma patients. Data collection was carried out over a three-month period from 15/05/2025 to 30/07/2025, at the Accidents and Emergency Department of Mulago National Referral Hospital (MNRH) in Kampala, Uganda. MNRH serves as the national tertiary referral and teaching hospital, receiving trauma casualties from across the country.

This analysis draws on the same cohort, study setting and ophthalmic examination protocol as our companion report on the prevalence and anatomical patterns of ocular injury in this population, posted as a preprint on medRxiv (**doi:** https://doi.org/10.64898/2026.08.06.26359857) and currently under review [13]. The present manuscript is restricted to identifying the socio-demographic and clinical correlates of ocular injury and does not duplicate the prevalence estimates or descriptive injury-pattern findings reported in the companion paper.

### Ethics statement

Ethical clearance for this study was granted by the School of Medicine Research and Ethics Committee (SOMREC) of Makerere University College of Health Sciences (Ref: Mak-SOMREC-2025-1254) and by the MNRH institutional review board; all study procedures conformed to the Declaration of Helsinki. Enrolled participants, or a legally authorized representative acting on their behalf, provided written informed consent prior to data collection. For unconscious patients with no caregiver available at the time of admission, enrolment proceeded under the committee’s waiver of initial consent, with deferred consent secured once the patient regained capacity or a caregiver could be traced.

### Participants

Consecutive adult patients (aged 18 years and above) admitted with a confirmed clinical diagnosis of head injury were enrolled into the study. Both conscious and unconscious patients were enrolled. A waiver of initial informed consent was obtained for unconscious patients without a primary caregiver; deferred consent was sought once the patient recovered or a caregiver was traced. Patients with pre-existing ocular disease unrelated to trauma were excluded.

### Sample size

The sample size for this analysis was calculated using Fleiss’ formula for comparing two independent proportions [14], based on the odds ratio for ocular injury among motor-vehicle passengers reported by Nalukenge et al. (AOR 3.85; 95% CI 1.49–9.93) [11]. Computations were performed in OpenEpi version 3 at 95% confidence, giving a minimum sample size of 223 participants; after a 10% adjustment for non-response, this rose to 246. Since the sample size for the study’s prevalence objective (383 participants) was larger, 383 was adopted as the final sample size.

### Data collection

A structured Kobo Toolbox questionnaire captured sociodemographic and clinical data for each participant. The principal investigator then performed a single standardized ophthalmic work-up on every patient, covering: distance visual acuity (3-metre Snellen or illiterate “E” chart), intraocular pressure measurement by calibrated Schiötz tonometry, anterior-segment evaluation by portable slit-lamp with fluorescein staining, pupillary light-reflex testing including the swinging-flashlight test for a relative afferent pupillary defect, assessment of ocular motility across the nine cardinal positions of gaze, indirect ophthalmoscopy with a Keeler Vantage Plus model for the dilated fundus, and confrontation visual fields where the patient could cooperate. Craniofacial computed tomography (CT) scans were subsequently reviewed for orbital, cranial and facial fractures and any accompanying soft-tissue injury.

### Outcome and variables

The primary outcome was the presence of at least one ocular injury, defined as any traumatic abnormality involving the ocular adnexa, eye globe, orbit, optic nerve, or cranial nerves supplying the ocular system. Eye globe injuries were classified using the Birmingham Eye Trauma Terminology (BETT) system. Head injury severity was graded using the Glasgow Coma Scale (GCS): mild (13-15), moderate (9-12), severe (8 and below). Independent variables included: age, sex, occupation, residence, mechanism of injury, GCS category, skull fractures (facial, cranial), intracranial haemorrhagic lesions, injury comorbidities, use of head protective equipment, and history of alcohol use.

### Statistical analysis

Analysis was performed in Stata 18.0 (StataCorp, College Station, TX). Continuous variables were presented as means with standard deviations, while categorical variables were presented as frequencies with percentages. The prevalence of ocular injury was calculated as the proportion of participants with at least one ocular injury, with a 95% confidence interval.

Bivariable modified Poisson regression with robust standard errors was used to generate crude prevalence ratios (CPR) with 95% confidence intervals for each independent variable. Predictors significant at p<0.2 on bivariable analysis were entered into a multivariable modified Poisson regression model to generate adjusted prevalence ratios (APR).

## Results

### Participant characteristics

We recruited 383 participants whose average age was 32.7 years of age (SD 12.3), and the cohort was predominantly male (89.3%) and urban-dwelling (65.5%). Road traffic accidents accounted for most injuries (68.9%), head injury was mild in 72.3% of cases, and documented use of head-protective equipment was uncommon (5.0%). Tables 1 and 2 give the complete socio-demographic and clinical profile of patients admitted with head injury.

**Table 1:** Socio-demographic characteristics of patients admitted with head injury at the Accidents and Emergency Unit of MNRH.

| Characteristic (N=383) | Frequency, n | Percentage % |
| --- | --- | --- |
| **Age group (years)** |  |  |
| 18- 25 (Young adults) | 133 | 34.7 |
| 26- 44 (Adults) | 190 | 49.6 |
| 45- 59 (Middle age) | 39 | 10.2 |
| ≥60 (Old age) | 21 | 5.5 |
| **Gender** |  |  |
| Female | 41 | 10.7 |
| Male | 342 | 89.3 |
| **Occupation** |  |  |
| Business person/ Self-employed | 105 | 27.4 |
| Motorcyclist (boda-boda rider) | 96 | 25.1 |
| Farmer | 73 | 19.1 |
| Casual laborer | 55 | 14.4 |
| Graduate employee | 30 | 7.8 |
| Others* | 24 | 6.2 |
| **Highest level of Education** |  |  |
| None | 24 | 6.3 |
| Primary | 205 | 53.5 |
| Secondary | 108 | 28.2 |
| Tertiary | 33 | 8.6 |
| University | 13 | 3.4 |
| **Residence** |  |  |
| Rural | 49 | 12.8 |
| Semi-urban | 83 | 21.7 |
| Urban | 251 | 65.5 |
| **Use of head protective equipment** |  |  |
| No | 179 | 46.7 |
| Yes | 19 | 5 |
| Unknown | 185 | 48.3 |
| **History of alcohol use at time of accident** |  |  |
| No | 326 | 85.1 |
| Yes | 22 | 5.7 |
| Unknown* | 35 | 9.1 |

*Occupation: Others\* Varied occupations that couldn't be captured under a unique category* *(these included being a; student, retired persons, witch doctor, security guard, housewife, parish* *chief, unknown patients). History of alcohol: Unknown\* Either the patient was unconscious, or* *their caretaker wasn't sure about history of alcohol use*

**Table 2:**
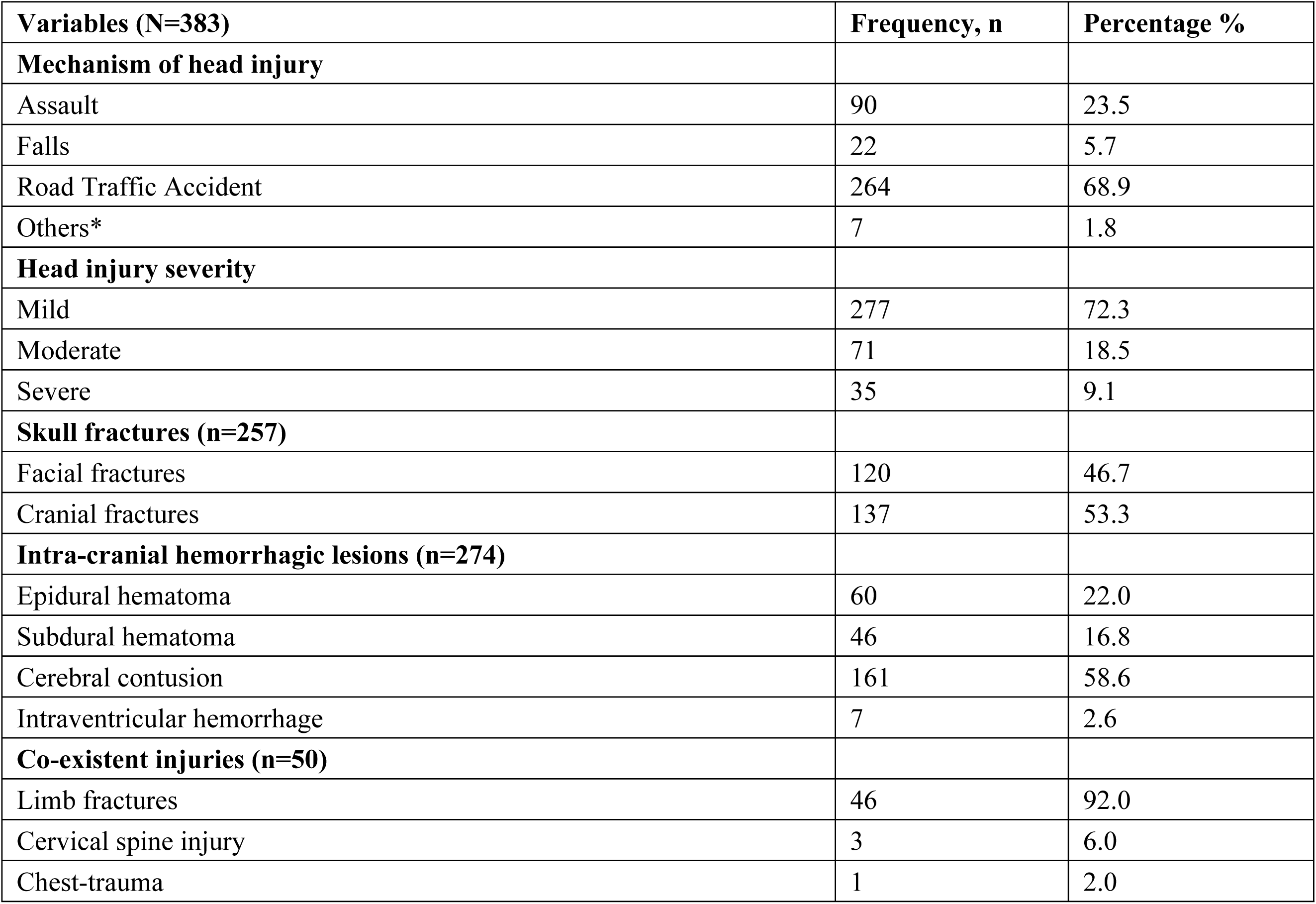
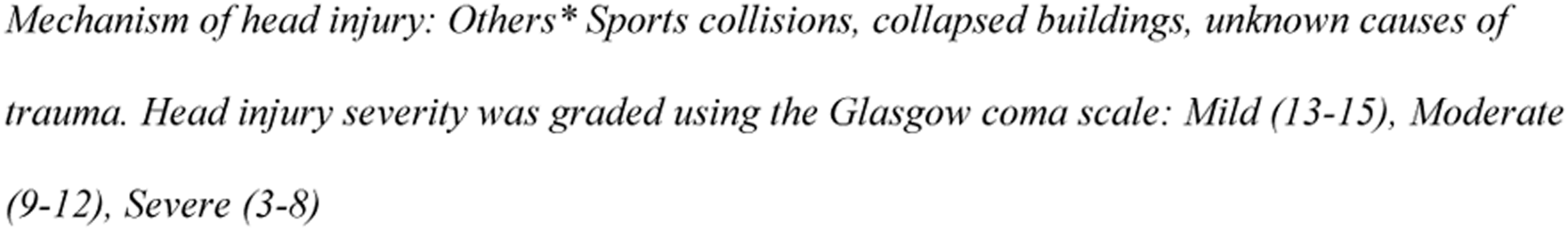
Clinical characteristics of patients admitted with head injury at the Accidents and Emergency Unit of MNRH.

| **Variables (N=383)** | **Frequency, n** | **Percentage %** |
| --- | --- | --- |
| **Mechanism of head injury** |  |  |
| Assault | 90 | 23.5 |
| Falls | 22 | 5.7 |
| Road Traffic Accident | 264 | 68.9 |
| Others* | 7 | 1.8 |
| **Head injury severity** |  |  |
| Mild | 277 | 72.3 |
| Moderate | 71 | 18.5 |
| Severe | 35 | 9.1 |
| **Skull fractures (n=257)** |  |  |
| Facial fractures | 120 | 46.7 |
| Cranial fractures | 137 | 53.3 |
| **Intra-cranial hemorrhagic lesions (n=274)** |  |  |
| Epidural hematoma | 60 | 22.0 |
| Subdural hematoma | 46 | 16.8 |
| Cerebral contusion | 161 | 58.6 |
| Intraventricular hemorrhage | 7 | 2.6 |
| **Co-existent injuries (n=50)** |  |  |
| Limb fractures | 46 | 92.0 |
| Cervical spine injury | 3 | 6.0 |
| Chest-trauma | 1 | 2.0 |

*Mechanism of head injury: Others\* Sports collisions, collapsed buildings, unknown causes of trauma. Head injury severity was graded using the Glasgow coma scale: Mild (13-15), Moderate (9-12), Severe (3-8)*

### Factors associated with ocular injury

Adults aged 26-44 years were 18% more likely to sustain ocular injury than those aged 18-25 years (APR 1.18; 95% CI 1.11-1.43; p=0.013). Commercial motorcyclists were 25% more likely to sustain ocular injury than business/ self-employed persons (APR 1.25; 95% CI 1.06-1.47; p=0.007). Road traffic accidents as the mechanism of head injury increased the likelihood of ocular injury by 19% compared to assault or falls (APR 1.19; 95% CI 1.02-1.38; p=0.03). Severe head injury increased the likelihood of ocular injury by 70% compared to mild head injury (APR 1.70; 95% CI 1.44-2.01; p<0.0001). The presence of facial fractures increased the likelihood of ocular injury by 61% (APR 1.61; 95% CI 1.44-1.81; p<0.0001). Intracranial haemorrhagic lesions were inversely associated with ocular injury (APR 0.80; 95% CI 0.68-0.93; p=0.004).

Details are presented in Table 3 and Figure 1.

**Figure 1.**
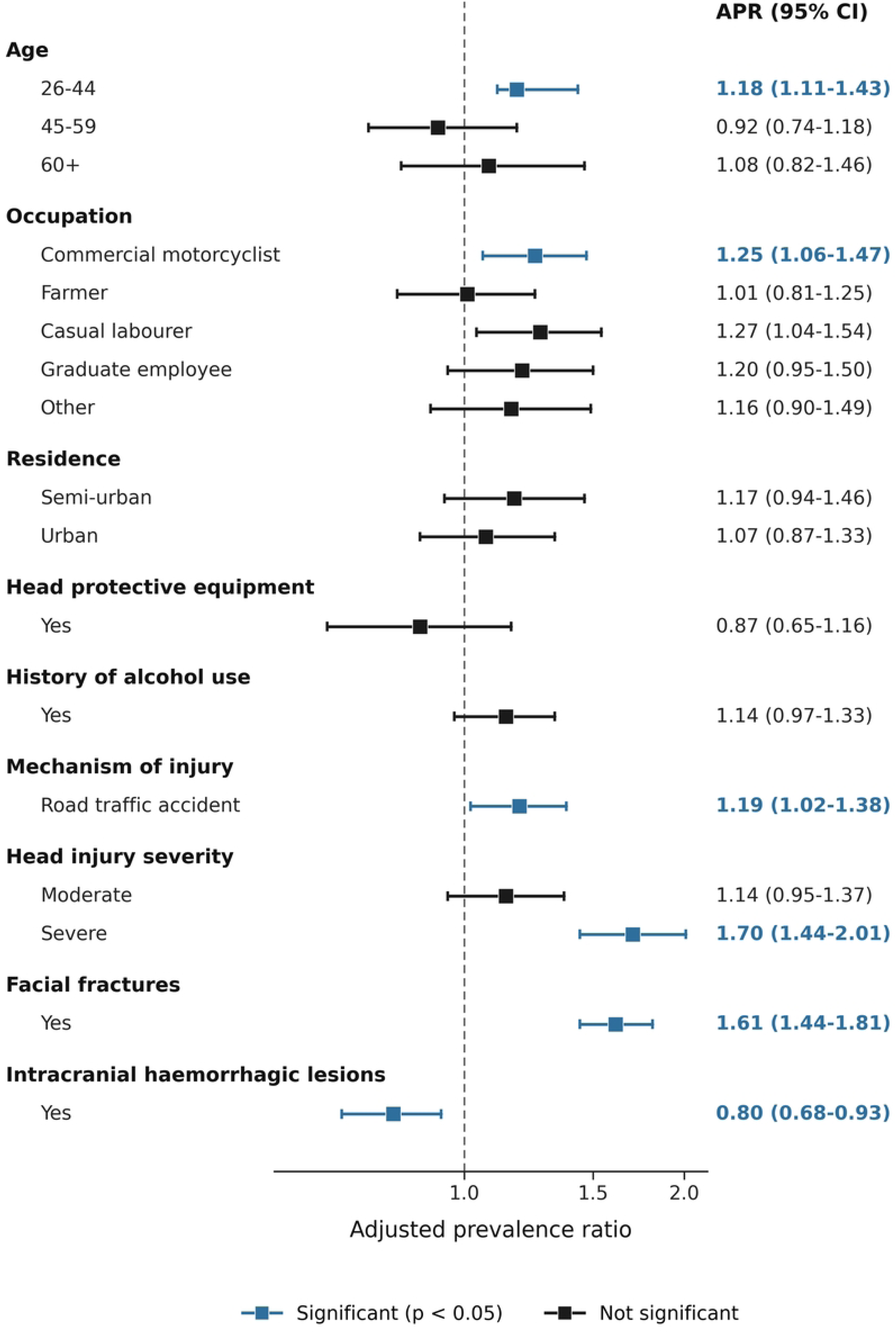
Forest plot of factors associated with ocular injury among patients admitted with head injury at the Accidents and Emergency Department of MNRH.

**Table 3:** Factors associated with ocular injury among patients admitted with head injury at the Accidents and Emergency Unit of MNRH.

| Variable | CPR (95% CI) | p-value | APR (95% CI) | p-value |
| --- | --- | --- | --- | --- |
| **Age (years)** |  |  |  |  |
| 18-25 (Young adults) | 1 |  | 1 |  |
| 26-44 (Adults) | 1.15 (1.00-1.33) | 0.06 | 1.18 (1.11-1.43) | 0.013 |
| 45-59 (Middle age) | 0.94 (0.71-1.24) | 0.63 | 0.92 (0.74-1.18) | 0.540 |
| ≥60 (Old age) | 1.02 (0.74-1.40) | 0.93 | 1.08 (0.82-1.46) | 0.610 |
| **Occupation** |  |  |  |  |
| Business person/ Self-employed | 1 |  | 1 |  |
| Commercial motorcyclist (boda-boda rider) | 1.27 (1.07-1.53) | 0.008 | 1.25 (1.06-1.47) | 0.007 |
| Farmer | 1.00 (0.79-1.26) | 0.97 | 1.01 (0.81-1.25) | 0.95 |
| Casual laborer | 1.20 (0.97-1.49) | 0.09 | 1.27 (1.04-1.54) | 0.71 |
| Graduate employee | 1.23 (0.97-1.59) | 0.09 | 1.20 (0.95-1.50) | 0.12 |
| Others* | 1.21 (0.92-1.60) | 0.17 | 1.16 (0.90-1.49) | 0.24 |
| **Residence** |  |  |  |  |
| Rural | 1 |  | 1 |  |
| Semi-urban | 1.28 (1.00-1.64) | 0.05 | 1.17 (0.94-1.46) | 0.16 |
| Urban | 1.13 (0.89-1.43) | 0.33 | 1.07 (0.87-1.33) | 0.51 |
| **Head protective equipment** |  |  |  |  |
| No/unknown | 1 |  | 1 |  |
| Yes | 0.98 (0.71-1.34) | 0.88 | 0.87 (0.65-1.16) | 0.34 |
| **History of alcohol use** |  |  |  |  |
| No/unknown | 1 |  | 1 |  |
| Yes | 1.09 (0.93-1.29) | 0.29 | 1.14 (0.97-1.33) | 0.11 |
| **Mechanism of injury** |  |  |  |  |
| Assault/falls | 1 |  | 1 |  |
| Road traffic accident | 1.25 (1.04-1.50) | 0.017 | 1.19 (1.02-1.38) | 0.030 |
| **Head injury severity** |  |  |  |  |
| Mild | 1 |  | 1 |  |
| Moderate | 1.04 (0.87-1.25) | 0.63 | 1.14 (0.95-1.37) | 0.170 |
| Severe | 1.51 (1.39-1.65) | <0.0001 | 1.70 (1.44-2.01) | <0.0001 |
| **Facial fractures** |  |  |  |  |
| No | 1 |  | 1 |  |
| Yes | 1.74 (1.56-1.93) | <0.0001 | 1.61 (1.44-1.81) | <0.0001 |
| **Intracranial haemorrhagic lesions** |  |  |  |  |
| No | 1 |  | 1 |  |
| Yes | 0.80 (0.71-0.91) | 0.001 | 0.80 (0.68-0.93) | 0.004 |

CPR: crude prevalence ratio; APR: adjusted prevalence ratio; CI: confidence interval. A
prevalence ratio of 1 indicates the reference category.

## Discussion

This study investigated factors associated with ocular injury among patients admitted with head injury at Mulago National Referral Hospital.

Ocular injury was independently associated with younger age, occupation as a commercial motorcyclist (boda-boda rider). Injury-related factors significantly associated with ocular injury included road traffic accidents as the mechanism of injury, severe head injury (low Glasgow coma scale score), presence of facial fractures, and intracranial hemorrhagic lesions.

These findings demonstrate that both socio-demographic characteristics and the severity and mechanism of trauma play a critical role in determining ocular involvement following head injury. The results are discussed below in relation to existing literature, and their clinical and public implications.

### Age

Patients aged 26 to 44 years were 18% more likely to sustain an ocular injury as compared to the younger and older patients. This can be explained by the fact that this adult age group constitutes the active work force of the country, they commonly engage in motorcycle riding, contact sports, and manual labor, consistent with studies from Uganda, Tanzania and Ethiopia, where young adult males dominate trauma admissions [11, 15–17].

### Occupation

Occupation has been shown to be an important determinant of ocular morbidity following head injury. Motorcyclists, particularly commercial riders (boda-boda riders), have consistently been reported to have higher risk of ocular and craniofacial injuries compared to other occupational groups. Our study showed a 25% higher likelihood for them to have ocular morbidity compared to other professions. Studies from Sub-Saharan Africa and other low-and middle-income countries demonstrate that motorcycle riders account for a disproportionally large portion of road traffic-related ocular trauma, largely due to frequent exposure to high-speed crashes, poor helmet use, and inadequate eye protection [11, 18, 19]. In Uganda, boda-boda riders have been identified as high-risk occupational group for head and ocular injuries, with poor helmet compliance and weak enforcement of road safety regulations contributing significantly to injury severity [20, 21]. Similar findings have been reported in studies from Nigeria and Tanzania, where motorcycle riders were more likely to sustain severe ocular injuries compared to occupants of other vehicle types [22, 23].

### Mechanism of head injury

Patients involved in road traffic accidents were more likely to sustain ocular injuries compared to those admitted with other mechanisms of injury. Road traffic accidents are a leading cause of head injury admissions globally, particularly in low-and middle-income countries, where they account for a substantial proportion of trauma-related morbidity and mortality [24–26]. The high energy forces involved in road traffic crashes, including rapid acceleration-deceleration, blunt impact, and rotational forces, increase the risk of both intracranial and ocular injuries [24, 27]. Globally, studies have demonstrated that ocular trauma commonly occurs in association with road traffic injuries and is an important contributor to visual impairment and blindness [7, 28]. These patterns have been consistently reported across different regions, including Europe, Asia and Africa, indicating that the association between road traffic accidents and ocular injury is not context-specific [7, 28]. In Uganda, *Kigera et al* similarly reported a higher likelihood of ocular injuries among patients involved in road traffic accidents, supporting the global evidence that road traffic trauma is a major risk factor for ocular morbidity [20].

### Severity of head injury

Patients with severe head injury were 70% more likely to sustain ocular trauma as compared to those with mild or moderate head injury. The severity of head injury is a critical determinant of the occurrence and pattern of ocular morbidity. Patients with severe head injury, typically defined by Glasgow Coma Scale (GCS) score of ≤8, consistently demonstrate a significantly higher prevalence of ocular injuries compared to those sustaining mild or moderate traumatic brain injury (TBI). A large-scale retrospective study by *Zhang et al* [29] found traumatic brain injury in nearly 58% of patients admitted with ocular trauma, with those suffering from severe TBI having 2.91 higher odds of sustaining permanent optic nerve or visual pathway injuries. This association is further corroborated by *Pattnaik et al* [30] whose prospective study of closed head injuries found a statistically significant association (p=0.02) between lower GCS and neurologically significant ocular signs such as relative afferent pupillary defect (RAPD).

Additionally, *Masila et al* in Kenya observed a clear positive correlation between severe head injury and severe ocular signs like papilledema [31]. The similarity across these studies reflects the common pathophysiological mechanism of energy transmission from high-velocity impact to orbital tissues. A possible reason for this association is that severe head trauma often involves high-impact mechanisms such as road traffic collisions, falls from height, or assaults, where both the skull and orbit absorb substantial energy. Clinically, this emphasizes the need for prompt ophthalmic evaluation in patients with low GCS, since ocular injuries may be masked by reduced consciousness.

### Facial fractures

Patients with facial fractures in this study were 61% as likely to sustain ocular injuries, a finding that is anatomically plausible given that the orbit forms an integral part of the midfacial skeleton and shares contiguous bony walls with the maxilla, zygoma, frontal, and nasal bones, facilitating transmission of traumatic forces to the orbital floor and walls [32].

Our findings are consistent with *Fomete et al* in Nigeria, who demonstrated that zygomaticomaxillary complex and naso-orbital-ethmoidal fractures were highly predictive of concomitant ocular injuries [33]. Although the literature shows variation in the type and severity of ocular injuries associated with different fracture patterns, the overall association midfacial fractures and ocular morbidity remains consistent. Large retrospective studies and systematic reviews report that orbital blowout and periorbital fractures are more frequently associated with ocular injuries overall, while zygomaticomaxillary complex fractures are more commonly linked to severe visual outcomes, including blindness, particularly following high-impact trauma [34–38].

Therefore, while the specific fracture pattern may influence the severity or type of ocular involvement, the cumulative evidence supports a strong and consistent relationship between facial fractures and ocular injury. Our study findings align with this body of literature and reinforce facial fractures as a significant predictor of ocular morbidity among patients with head trauma.

### Intracranial hemorrhagic lesions

Interestingly, patients in our study who presented with intracranial hemorrhagic lesions had a 20% lower probability of sustaining ocular injuries compared to those without such lesions. This inverse association was unexpected, as both intracranial and orbital injuries often result from high-energy trauma. One possible explanation relates to differences in injury biomechanics rather than a true protective effect. The pattern and distribution of traumatic force influence whether impact energy is dissipated within the cranial vault or transmitted to the orbital structures and causing concurrent ocular trauma. *Kato et al* demonstrated that intracranial pathology correlates with mechanism and direction of impact, which may determine the extent of associated craniofacial involvement [39]. Similarly, *Snyder et al* reported that certain intracranial hemorrhagic patterns, particularly isolated intracranial hemorrhage without skull or facial fracture, were associated with lower odds of ocular (specifically retinal) hemorrhage [40]. *Zhang et al* further emphasized that not all categories of traumatic brain injury were accompanied with ocular injury, suggesting that the transmission pathway and absorption of force determine whether the orbit is affected [29].

These findings suggest that intracranial hemorrhage and ocular injuries do not always occur together and may result from differences in how the force of trauma is distributed. Therefore, the inverse association observed in our study should be interpreted cautiously and does not imply that intracranial hemorrhage is protective against ocular injury.

### Strengths and limitations

A major strength of this study was the inclusion of both conscious and unconscious patients rather than limiting enrolment to those able to cooperate with full examination, thereby minimizing selection bias. Furthermore, applying a uniform, detailed ophthalmic protocol; including tonometry and comprehensive anterior and posterior segment evaluations to every participant regardless of trauma severity ensured that risk factor estimates were not confounded by variable case ascertainment.

However, several limitations warrant consideration when interpreting these findings. Since recruitment was conducted at a single tertiary referral center handling a high volume of severe trauma, strong associations-particularly those involving severe head injury and facial fractures, may not fully generalize to populations treated in primary level healthcare facilities.

Additionally, exposure data for variables such as helmet usage and alcohol intake relied on patient or caregiver reports, introducing potential recall and reporting bias. Due to ocular assessments being conducted solely at admission, ophthalmic complications that evolved later during clinical recovery were not captured in this baseline snapshot. Finally, the cross-sectional design demonstrates statistical associations between identified risk factors and ocular injury, but precludes establishing causality.

## Conclusion

Ocular injuries are common among patients with head injury. Patients aged 26 to 44 years, commercial motorcyclist occupation, road traffic injury, severe head injury, and facial fractures have a higher risk of ocular injury. We recommend prioritizing ophthalmic evaluation of for patients admitted with head injury to minimize preventable visual impairment, particularly among patients presenting with; severe head injury, facial fractures, or road-traffic-related trauma, as well as commercial motorcyclists.

## Data Availability

The data set used in this manuscript have been attached in the supporting files.

## Acknowledgments

The authors acknowledge the patients and their caregivers who consented to participate in this study, the staff of the Accidents and Emergency Department of MNRH, and the research assistants for their contribution to the collection of data.

## Supporting information

S1 Data set for patients admitted with head injury at MNRH

